# Material deprivation is associated with neural resilience for late-life depression

**DOI:** 10.1101/2022.07.25.22277997

**Authors:** Wei Zhang, Janine D. Bijsterbosch

**Affiliations:** Department of Radiology, Washington University School of Medicine, St. Louis, Washington University School of Medicine, St. Louis

**Keywords:** late-life depression, material deprivation, neural resilience, neuroanatomical signatures

## Abstract

Late-life depression (LLD) is a major source of global morbidity and mortality, influenced by multiple risk factors. Yet, a major challenge is to quantify the degree of resilience or vulnerability to LLD at the individual level, which could offer neurobiological insight and ultimately inform future interventions and treatment. Here, applying a non-parametric regression model to the UK Biobank data (N=1,988), we quantified brain-based resilience and vulnerability to LLD and tested whether risk factors could explain individual differences in the estimated magnitude of such neural resilience and vulnerability. Our results show that social isolation was positively associated with the median magnitude of neural vulnerability whereas material deprivation was negatively associated with the greatest neural resilience (top 10 percentile). These results together highlight the importance of social interaction and access to sufficient resources and services in diminishing neural vulnerability and promoting neural resilience to LLD, respectively. Our findings therefore provide insights into preventive strategies for LLD, and thus are of importance for policy makers as well as the broader society.

## Introduction

Late life depression (i.e., depression in older adults aged 60+; LLD) has been associated with increased risks of disability and mortality (Schulz *et al*., 2000). It is intertwined with conditions and factors that are primarily ageing-related, yielding distinctive and complex etiological and clinical profiles in contrast to depression in younger age groups (Blazer, 2003; Alexopoulos, Schultz and Lebowitz, 2005). Previous work has shown that risk factors such as physical disability, medical illness, cognitive impairment, worse socioeconomic status, greater exposures to traumatic events, less social support, and living an unhealthy lifestyle contribute to higher chances of depression (Blazer, 2003; Chang-Quan *et al*., 2010; Chang *et al*., 2016). However, these studies did not investigate individual variation in LLD vulnerability and/or resilience and how such variation may be linked to various risk factors. Quantifying LLD resilience and vulnerability in patients is of great clinical relevance as it may ultimately inform future interventions. Here we employed a novel approach to evaluate neural resilience and vulnerability to LLD at the individual level and determined whether known risk factors of LLD explain individual differences in such neural resilience and vulnerability.

In previous studies, LLD has been associated with abnormalities in structural and functional properties (Manning and Steffens, 2018). It has also been associated with accelerated brain age, the magnitude of which further showed a correlation with declined cognitive performance (Christman *et al*., 2020). These findings demonstrate the potential of proxy measures that capture the neural underpinnings of the disorder. Here, we build on this prior work to assess resilience and vulnerability to depressive symptom severity, using similar aggregate neural measures. Specifically, we sought to quantify such resilience and vulnerability from neuroanatomical patterns that are related to recent depressive symptoms and to examine whether LLD risk factors could explain individual differences in the quantified neural resilience and vulnerability. Specifially, here we focused on a vulnerable group of older adults who have experienced at least one depression episode before the assessment in this study. This provides an opportunity to enhance the variance in depression symptom severity by including individuals along the spectrum of potential LLD. Inspired by brain-age models that predict the age of the brain based on neuroanatomical features in healthy individuals (Franke and Gaser, 2019), here we predicted the individual-level brain-based depression score (BDS) from multimodal neuroimaging features (n=4,632; see more details in *Materials and Methods*). The difference between the predicted BDS and the original symptom score (delta BDS; Δ*BDS)* was calculated to index “neural resilience” or “neural vulnerability” to LLD and then linked to a set of LLD risk factors that covered a wide range of sociopsychological, medical and lifestyle variables (see variable full list in *Materials and Methods*). We specifically focused on individuals with highest neural resilience or vulnerability (i.e., top 10% negative and positive Δ*BDS* respectively) that represent older adults with the greatest clinical relevance. In practice, the focus on these highly resilient and vulnerable individuals also minimized the correlations between Δ*BDS* and original symptom score (i.e., similar to correlations between brain-age delta and chronological age in brain-age literature; (Franke and Gaser, 2019; Smith *et al*., 2019). This enabled us to identify risk factors showing unique associations with neural resilience and/or vulnerability.

## Materials and Methods

### Participants

Participants from the UK Biobank (UKB) dataset aged 60 or above at the time of imaging acquisition with a history of probable major depression status (i.e., prior experiences in single or recurrent depression episodes; (Smith *et al*., 2013)) were included to ensure sufficient variance in depression symptomatology (M_age_=66.35; 60.3% female). Approximately 70% (N=1,405) of the resulting sample (N=1,988) had a status of recurrent major depression. Due to missing data in risk factor variables, a sub-sample of N=1,464 participants were included in partial correlation and quantile regression analyses with similar demographic characteristics and prior depression histories as in the full sample (M_age_=66.18; 59.4% female; 70% recurrent MDD). All participants in this study provided informed consent. UK Biobank has ethical approval from the North West Multi-Centre Research Ethics Committee (MREC). Data access was obtained under UK Biobank application ID 47267.

### Data acquisition and measurement

#### Depressive symptoms

A touchscreen questionnaire was implemented to collect information on sociodemographic characteristics and mental health. Recent depressive symptoms were measured using four items (RDS-4) that assess depressed mood, disinterest, restlessness, and tiredness in the past two weeks (Dutt *et al*., 2022). This continuous measure is highly comparable to several standardized self-report depression scales including PHQ-9, CES-D, and MASQ-30 (Dutt *et al*., 2022), and shows an area under the curve of 0.79 for its correlation with depression diagnosis (Khubchandani *et al*., 2016). In response to each of these four questions, participants indicated their experiences from “not at all” (scoring 1) to “nearly every day” (scoring 4) such that the total symptom score ranged from 4 to 16.

#### Imaging preprocessing and multimodal imaging features

All brain imaging data from the UKB were acquired using standard Siemens Skyra 3T scanners with a standard Siemens 32-channel RF receive head coil. Detailed information for UKB imaging acquisition can be found in the UK Biobank Imaging Documentation, hosted on the Oxford FMRIB UK Biobank resource page (https://www.fmrib.ox.ac.uk/ukbiobank/). A fully automated processing and QC pipeline was developed for UKB brain imaging data, which included T1, T2, FLAIR, susceptibility-weighted MRI, resting-state MRI, task-evoked MRI and diffusion MRI (Alfaro-Almagro *et al*., 2018). Additionally, this pipeline also generated a set of imaging-derived phenotypes (IDPs) such as cortical and subcortical structure volumes, microstructural measures in major tracts, and functional connectivity metrics. In this study, a total of 4,632 IDPs was included as multimodal imaging features in the prediction models for estimating brain-depression score (see section *3*.*1*). Approximately three quarters of these features were derived from the resting-state and task-evoked fMRI data, and the remaining from structural and diffusion MRI data:

– Resting-state fMRI features: Amplitude of ICA100 nodes and ICA25 nodes; Edges of full correlation matrix from ICA100 and ICA25; Edges of partial correlation matrix from ICA100
– Task-evoked fMRI features: Median and 90th percentile BOLD signals for shapes and faces, as well as shape-face contrasts using a group mask and an amygdala mask respectively; Median and 90th percentile Z-statistics for shapes and faces, as well as shape-face contrasts using a group mask and an amygdala mask respectively
– Structural MRI features: Volumes of cortical and subcortical (including sub-segments) structures; Total volume of white matter hyperintensities
– Diffusion MRI features: Mean FA, MD, MO, L1-L3, ICVF, OD, ISOVF based on Standardized FA Skeleton; Weighted-mean FA, MD, MO, L1-L3, ICVF, OD, ISOVF in White-matter tracts

#### Late-life Depression Risk factors

A list of risk factors for late-life depression (LLD) was curated based on the literature that covers information on demographics, lifestyle, medical conditions, adverse experiences, and psychosocial factors. Measures of these variables were collected from participants either via a touchscreen questionnaire or a verbal medical history interview on the same day of imaging acquisition. Specifically, several variables such as self-reported health, long-standing illness and Townsend deprivation index, which is a census-based index of material deprivation calculated by the combination of four indicators of deprivation: non-home ownership, non-car ownership, unemployment and overcrowding (Townsend, Phillimore and Beattie, 1988) were available directly from the UKB, whereas aggregate measures for healthy lifestyle, sleep quality, vascular risk factors, adverse or traumatic experiences, social isolation and loneliness were derived using multiple items as follows:

– A *healthy lifestyle score* was constructed based on smoking status, physical activity, diet, and alcohol consumption that are well documented as depression risk factors (Sarris *et al*., 2020; van Lee *et al*., 2020; Kang *et al*., 2021). Based on national recommendations, participants were given 1 for healthy, and 0 for unhealthy behaviors. Detailed coding information can be found in (Lourida *et al*., 2019).
– A *sleep (low) risk score* was calculated using five sleep questions. Low-risk sleep factors were defined as a) having an early chronotype, b) sleeping 7–8 hours per day, c) never having or rarely having insomnia symptoms, d) not reporting snoring, and e) not reporting frequent daytime sleepiness (Fan *et al*., 2020). Participants received a score of 1 if their behaviors were classified as low risk for that factor and a sum score was calculated across five factors, where higher scores represent healthier sleep patterns or low risks of sleep issues (Hepsomali and Groeger, 2021).
– An *aggregate measure of vascular risk factors* was calculated for each participant by counting instances of having a high BMI (>25), having a high waist-hip ratio (WHR>0.85 for females and WHR>0.90 for males), having ever smoked, and a self-reported diagnosis of hypertension, diabetes, or hypercholesterolemia (Cox *et al*., 2019). The resulting sum score of instances represent potential vascular risks, where higher scores indicating higher risks.
– *scores* were calculated separately for childhood, adulthood, and lifetime experiences with dichotomization of responses to each individual question (Yapp *et al*., 2021). Specifically, responses of “never true” to negative experiences (e.g., hit hard) scored a 0 and responses of “rarely true” and more often (i.e., “sometimes true”, “often”, “very often true”) scored a 1, which was reversed for positive experiences (e.g., in a confiding relationship). Binary responses (“yes”, “no”) were scored 1 and 0 respectively. Separate sum scores were calculated to indicate the magnitude of traumatic experiences in different time periods and included as separate predictors in the models.
– *Psychosocial factors* comprised two measures: loneliness and social isolation. Participants were classified as lonely if they reported feeling lonely often and if they could confide to someone close only occasionally (e.g., less than once every few months), and socially isolated if they met at least two criteria of 1) living alone, 2) visiting their family or friends less than once a month, and 3) participating in none of the listed leisure/social activities (Mutz, Roscoe and Lewis, 2021). These psychosocial factors were included in statistical models as separate predictors.

### Statistical analysis

#### Estimating the delta of brain-depression score

Our approach was inspired by the estimation of brain-age delta from neuroimaging features that is defined as the difference between the estimated brain age and chronological age in a given individual, which has been used to indicate underlying problems in outwardly healthy people and related to the risk of cognitive ageing or age-associated brain disease (Franke *et al*., 2010; Cole and Franke, 2017; Baecker *et al*., 2021). In this study, we employed Multivariate Adaptive Regression Splines models (Friedman and Roosen, 1995; Friedman, 2007) to estimate the brain-depression score (BDS) as informed by multimodal imaging features and quantified the difference between the estimated and the actual depression symptom scores (i.e., Δ*BDS*), with a positive Δ*BDS* indicating neural vulnerability to depression (i.e., actual reported symptom score higher than BDS) and negative Δ*BDS* indicating neural resilience to depression (i.e., actual symptom score lower than BDS).

Multivariate Adaptive Regression Splines (MARS) is a flexible regression technique that can capture the intrinsic nonlinear and multidimensional relationship of variables with an ensemble of linear functions joined together by one or more spline basis functions, where the number of basis functions and the parameters associated with each function (e.g., product degree and knot locations) are determined by the data (Friedman and Roosen, 1995; Friedman, 2007). Specifically, MARS builds a model in two phases: the forward pass and the backward pruning, similar to growing and pruning of tree models. In the forward pass, MARS starts with a model consisting of just the intercept term (i.e., the mean of the response values), followed by the assessment of every single predictor to find a basis function pair that produces the maximum improvement in the model error. This process iterates until either the model reaches a predefined limit number of terms, or the error improvement reaches a predefined limit. The result of the forward pass is a MARS basis matrix with rows of observations and columns of basis functions (e.g., hinge functions). To avoid overfitting by the full terms in the basis matrix from the forward pass, the backward pass is to find the subset of these terms that gives the best generalized cross validation (GCV) via a stepwise term deletion procedure. This backward pruning process continues until only one term remains (the intercept term) and the final model with the best GCV is selected (Friedman and Roosen, 1995; Friedman, 2007).

In this study, IDPs from multimodal imaging data and depressive symptom sum scores were included in MARS models as independent (*X*) and dependent (*Y*) variables. Additionally, age, sex, head size, head motion (i.e., the averaged head motion across space and time points) during fMRI acquisition (i.e., for both the resting-state and task-evoked fMRI), scanner site, scanner table position and data acquisition dates were included in all MARS models as confounding variables. Per partition of the full data in the nested cross-validations (see details below), principal component analysis (PCA) was employed to decompose high-dimensional *X* and components collectively explaining more than 50% variance were retained. As the outcome measure *Y* (i.e., symptom sum score) in our study is highly skewed with a long tail, transformation of *Y* was performed per data partition before modeling to enforce Gaussianity. This was realized by using a data-drive approach that finds the optimal normalization method from a suite of possible transformation options including the Box-Cox transformation, Yeo-Johnson transformation, the ordered quantile technique, Arcsinh transformation, exponential transformation, square root transformation and the Lambert W x F transformation (Peterson, 2021). We also applied dummy coding for all factorial variables such as sex and scanner site before data partitioning to ensure the same number of predictors across models.

Nested cross-validations were applied to increase robustness and generalizability of our MARS estimations, with 3 iterations in the outer loop and 10 folds per iteration in the inner loop. As mentioned earlier, data processing including PCA (on *X*) and transformation (on *Y*) was performed within each iteration of the outer loop and parameters obtained from the training and validating set were applied to the held-out testing set. Per training fold, a grid search was performed to identify the optimal combination of two parameters: the maximum degree of interactions among terms (*ndegree*) and the number of terms retained in the final model (*nprune*). Given the available data points and the empirical evidence that 3^rd^-degree interactions never benefited model fit using a subset of data, expansion of interaction degree was restricted to 2 (e.g., testing *ndegree* = 1 and = 2) and *nprune* up to 5 times of the predictor numbers (i.e., varied between 690 and 695 due to changing numbers of principal components in each fold). Per prediction model on the testing set, we further partitioned data to estimate prediction uncertainty as the noise and model variance of the out-of-fold predicted values over 5 iterations of 50 cross-validations, using variance models of MARS (Milborrow, 2015), and an averaged r-squared of 0.19 in contrast to 0.09 from the training models.

For each participant, we quantified Δ*BDS* as the difference between the predicted BDS (*ŷ*) and the reported symptom sum score (*Y*) while accounting for the prediction uncertainty including irreducible “aleatoric” or noise variance 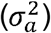 and model variance of prediction or “epistemic” uncertainty 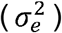 such that 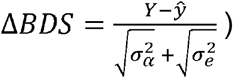. By definition, the resulting Δ*BDS* for each participant indexed less depressed or resilient brain patterns if the quantity was negative, and more depressed or vulnerable brain patterns if positive. Here, we focused on individuals with high neural resilience (i.e., top 10 percentile of the negative Δ*BDS*) and individuals with high neural vulnerability (i.e., top 10 percentile of the positive Δ*BDS*). This was determined to drive a good balance between sufficient statistical power (i.e., n= 144 with 11 regressors) and high clinical relevance, and to reduce dependency of BDS as a function of original symptom score (i.e., minimize high correlation between Δ*BDS* and original symptom score).

#### Testing risk factor effects

All analyses were conducted for individuals with high neural resilience and vulnerability separately. We first conducted partial rank correlation analyses to test the associations between each risk factor and the Δ*BDS* while controlling for the reported symptom sum score (i.e., resulting in effects not influenced by the sum score). This is a common practice to deconfound the associative effects driven by chronological age rather than the brain-age delta in the brain age literature (Smith *et al*., 2019). False Discover Rate (FDR) corrections were applied to adjust for multiple testing. We further conducted quantile regression analyses to identify unique contributions of individual risk factors, while accounting for the effects of reported symptom sum score.

## Results

For each participant, we estimated the BDS using the Multivariate Adaptive Regression Splines (MARS) model (*Figure 1*). Our model yielded successful prediction of the BDS from the held-out sample via nested cross validations (RMSE=0.90, R^2^=0.09), with a comparable effect size to recent brain-wide association studies that had sufficiently large sample sizes (Dick *et al*., 2021; Marek *et al*., 2022). Importantly, our MARS model captures information about brain structural and functional features associated with LLD symptoms and thus indicates the disorder manifestation at the neurobiological level (i.e., a proxy of how “depressed” an individual’s brain is).

**Figure 1.**
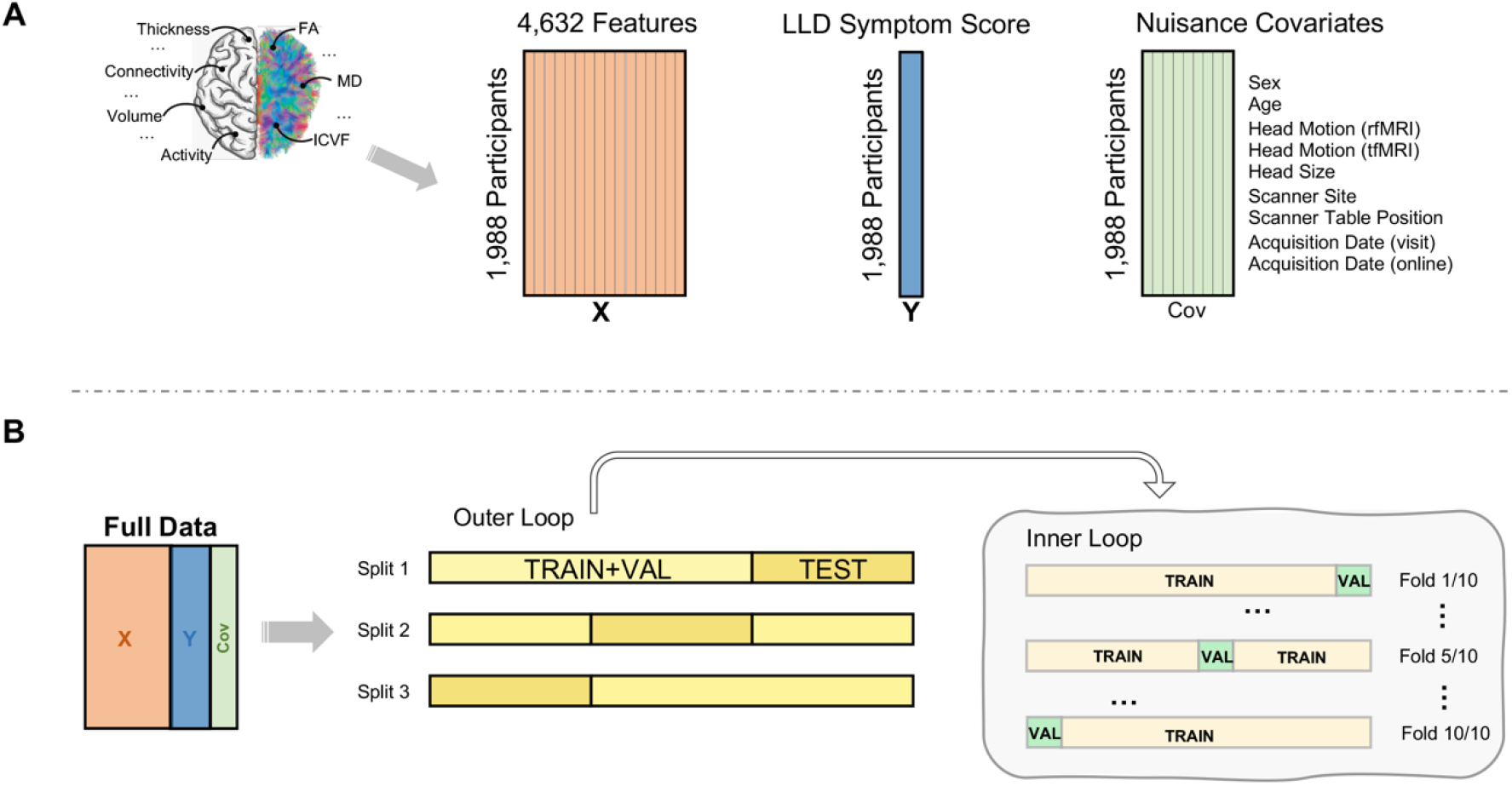
Prediction Models for Brain-depression Score. (A) Multimodal imaging features including cortical thickness, functional connectivity, gray matter volume, task activity and white matter microstructure measures including fractional anisotropy (FA), mean diffusivity (MD) and intracellular volume fraction (ICVF), as well as late-life depressive (LLD) symptom score and nuisance covariates were included into multivariate adaptive regression splines models. (B) In each model, multimodal imaging features (*X*) were used to predict symptom sum score (*Y*) while controlling for sex, age, in-scanner motion, head size, scanner site, scanner table position and data acquisition dates (*Cov*), via nested cross-validations. In the outer loop, full data were partitioned into 3 splits and in the inner loop, each split of training and validating set (TRAIN+VAL) was fed into a 10-fold cross-validation to validate the model.

In this study sample, about 58% (n=1,144) participants appeared to possess a resilient brain as their BDS was lower than their reported depressive symptom scores (i.e., negative Δ*BDS*), and 42% showed the opposite pattern with a vulnerable brain (i.e., positive Δ*BDS*). Results from partial correlation analyses show that the Townsend deprivation index was negatively correlated with the magnitude of neural resilience after FDR corrections (r=-0.26, FDR corrected p=0.018). Findings from quantile regression analyses further show that this effect is mostly robust in participants with the highest level of neural resilience (e.g., top 10 percentile with the largest negative Δ*BDS*) when all risk factors were considered in one model (t=3.72, p<0.001; *Figure 2*). Interestingly, this association with the material deprivation was not observed for individuals with the highest neural vulnerability (i.e., top 10 percentile with the largest positive Δ*BDS*).

**Figure 2.**
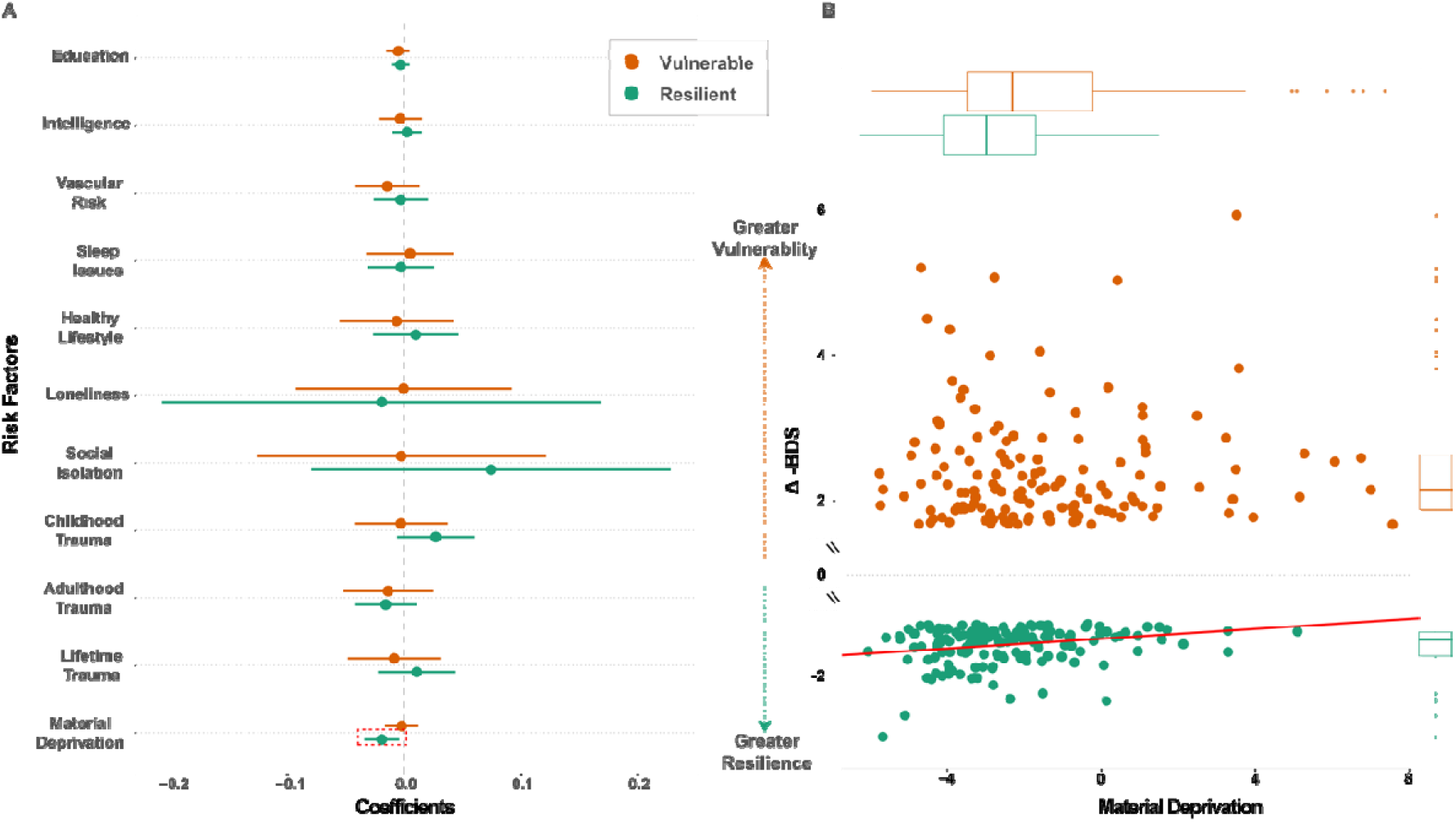
Impact of Risk factors on Brain-depression Index. (A) The dot-and-whisker plot compares the estimated coefficient and its standard error for each risk factor from separate multiple linear regression models for high neural resilience and vulnerability. (B) Material deprivation was significantly associated with the delta of brain-depression score (Δ*BDS*) only for individuals with high neural resilience.

Neither did we observe this associative effect for the self-reported depressive symptom scores from individuals with the most resilient or vulnerable neural patterns. These findings together suggested a specific effect of material deprivation on the LLD-related neural resilience. Additionally, we observed a positive association between social isolation and the median quantile magnitude of neural vulnerability with adjustment for all other risk factors (t=2.01, p<0.05). This associative effect was absent in participants showing resilient patterns at the neuroanatomical level.

## Discussion

In this study, we quantified individual-specific neural resilience and vulnerability to LLD using multimodal imaging features and linked known risk factors of LLD to such neural resilience and vulnerability. Our findings provide empirical evidence that risk factors may exert varying impact on the neurobiological manifestation of LLD. Specifically, we found a negative association between material deprivation and the Δ*BDS* in individuals with high neural resilience. This result indicated a beneficial effect of sufficient material resources on neural resilience to LLD. Interestingly, on the other hand, more material deprivation was not associated with neural vulnerability, suggesting that insufficient material resources alone may not adequately contribute to the differential patterns of brain structure and function that put individuals at increased risks of depression. Additionally, we observed a positive association between neural vulnerability to LLD and social isolation, which was absent in participants showing neuroanatomically resilient patterns. These findings are consistent with the observations that older adults with sufficient resources to age successfully are relatively healthy, active, independent and maintain high levels of mental well-being (Terraneo, 2021), and that increases in socioeconomic status decreases the odds for depression (Freeman *et al*., 2016; Zhou *et al*., 2021). Furthermore, our results also resonate with previous findings that social isolation and/or loneliness in older adults is associated with increased risk of all-cause mortality (Holt-Lunstad *et al*., 2015), as well as clinically significant depression and anxiety (Schwarzbach *et al*., 2014; Taylor *et al*., 2018; Domènech-Abella *et al*., 2019; Donovan and Blazer, 2020). It is however, important to note that the BDS was estimated from the symptom sum score in the current study and thus might fail to capture subtleties in neuroanatomical features associated with specific clinical subtypes. It is highly likely that the disorder manifests in a heterogenous manner and the presentation of vulnerability at the neuroanatomical level can vary across individuals. Future studies may consider using individual symptom-level or dimensional measures of LLD to estimate BDS and test risk factor effects on those potential subtypes.

In conclusion, our results demonstrate a link between material deprivation and neural resilience, as well as between social isolation and neural vulnerability to LLD. These results are of importance for policy makers as well as the broader society, as they provide evidence that sufficient material resources can improve neural resilience to depression for older adults and that compensational solutions to improve human interactions are in urgent need to offset the potential vulnerability to LLD when in-person social contacts are restricted.

## Data Availability

Data used in this study are publicly available at UK Biobank (https://www.ukbiobank.ac.uk/). All data produced in the present study are available upon reasonable request to the authors.

## Acknowledgments

This research was performed under UK Biobank application number 47267. This research was supported by the NIH (1 R34 NS118618-01) and the McDonnell Center for Systems Neuroscience.

## References

Alexopoulos, G. S., Schultz, S. K. and Lebowitz, B. D. (2005) ‘Late-Life Depression: A Model for Medical Classification’, Biological Psychiatry. Elsevier, 58(4), p. 283. doi: 10.1016/J.BIOPSYCH.2005.04.055.

Alfaro-Almagro, F. et al. (2018) ‘Image processing and Quality Control for the first 10,000 brain imaging datasets from UK Biobank’, NeuroImage. doi: 10.1016/j.neuroimage.2017.10.034.

Baecker, L. et al. (2021) ‘Machine learning for brain age prediction: Introduction to methods and clinical applications’, EBioMedicine. doi: 10.1016/j.ebiom.2021.103600.

Blazer, D. G. (2003) ‘Depression in Late Life: Review and Commentary’, The Journals of Gerontology: Series A. Oxford Academic, 58(3), pp. M249–M265. doi: 10.1093/GERONA/58.3.M249.

Chang-Quan, H. et al. (2010) ‘Education and risk for late life depression: A meta-analysis of published literature’, International Journal of Psychiatry in Medicine. doi: 10.2190/PM.40.1.i.

Chang, S. C. et al. (2016) ‘Risk factors for late-life depression: A prospective cohort study among older women’, Preventive Medicine. doi: 10.1016/j.ypmed.2016.08.014.

Christman, S. et al. (2020) ‘Accelerated brain aging predicts impaired cognitive performance and greater disability in geriatric but not midlife adult depression’, Translational Psychiatry. doi: 10.1038/s41398-020-01004-z.

Cole, J. H. and Franke, K. (2017) ‘Predicting Age Using Neuroimaging: Innovative Brain Ageing Biomarkers’, Trends in Neurosciences. doi: 10.1016/j.tins.2017.10.001.

Cox, S. R. et al. (2019) ‘Associations between vascular risk factors and brain MRI indices in UK Biobank’, European Heart Journal. doi: 10.1093/eurheartj/ehz100.

Dick, A. S. et al. (2021) ‘Meaningful associations in the adolescent brain cognitive development study’, NeuroImage. doi: 10.1016/j.neuroimage.2021.118262.

Domènech-Abella, J. et al. (2019) ‘Anxiety, depression, loneliness and social network in the elderly: Longitudinal associations from The Irish Longitudinal Study on Ageing (TILDA)’, Journal of Affective Disorders. doi: 10.1016/j.jad.2018.12.043.

Donovan, N. J. and Blazer, D. (2020) ‘Social Isolation and Loneliness in Older Adults: Review and Commentary of a National Academies Report’, American Journal of Geriatric Psychiatry. doi: 10.1016/j.jagp.2020.08.005.

Dutt, R. K. et al. (2022) ‘Mental health in the UK Biobank: A roadmap to self-report measures and neuroimaging correlates’, Human Brain Mapping. doi: 10.1002/hbm.25690.

Fan, M. et al. (2020) ‘Sleep patterns, genetic susceptibility, and incident cardiovascular disease: A prospective study of 385 292 UK biobank participants’, European Heart Journal. doi: 10.1093/eurheartj/ehz849.

Franke, K. et al. (2010) ‘Estimating the age of healthy subjects from T1-weighted MRI scans using kernel methods: Exploring the influence of various parameters’, NeuroImage. doi: 10.1016/j.neuroimage.2010.01.005.

Franke, K. and Gaser, C. (2019) ‘Ten years of brainage as a neuroimaging biomarker of brain aging: What insights have we gained?’, Frontiers in Neurology. doi: 10.3389/fneur.2019.00789.

Freeman, A. et al. (2016) ‘The role of socio-economic status in depression: Results from the COURAGE (aging survey in Europe)’, BMC Public Health. doi: 10.1186/s12889-016-3638-0.

Friedman, J. H. (2007) ‘Multivariate Adaptive Regression Splines’, The Annals of Statistics. doi: 10.1214/aos/1176347963.

Friedman, J. H. and Roosen, C. B. (1995) ‘An introduction to multivariate adaptive regression splines’, Statistical Methods in Medical Research. doi: 10.1177/096228029500400303.

Hepsomali, P. and Groeger, J. A. (2021) ‘Diet, sleep, and mental health: Insights from the uk biobank study’, Nutrients. doi: 10.3390/nu13082573.

Holt-Lunstad, J. et al. (2015) ‘Loneliness and Social Isolation as Risk Factors for Mortality: A Meta-Analytic Review’, Perspectives on Psychological Science. doi: 10.1177/1745691614568352.

Kang, M. et al. (2021) ‘The relationship of lifestyle risk factors and depression in Korean adults: A moderating effect of overall nutritional adequacy’, Nutrients. doi: 10.3390/nu13082626.

Khubchandani, J. et al. (2016) ‘The Psychometric Properties of PHQ-4 Depression and Anxiety Screening Scale Among College Students’, Archives of Psychiatric Nursing. doi: 10.1016/j.apnu.2016.01.014.

van Lee, L. et al. (2020) ‘Multiple modifiable lifestyle factors and the risk of perinatal depression during pregnancy: Findings from the GUSTO cohort’, Comprehensive Psychiatry. doi: 10.1016/j.comppsych.2020.152210.

Lourida, I. et al. (2019) ‘Association of Lifestyle and Genetic Risk With Incidence of Dementia’, JAMA, 322(5), p. 430. doi: 10.1001/jama.2019.9879.

Manning, K. J. and Steffens, D. C. (2018) ‘State of the Science of Neural Systems in Late-Life Depression: Impact on Clinical Presentation and Treatment Outcome’, Journal of the American Geriatrics Society. NIH Public Access, 66(Suppl 1), p. S17. doi: 10.1111/JGS.15353.

Marek, S. et al. (2022) ‘Reproducible brain-wide association studies require thousands of individuals’, Nature. Springer US, 603(7902), pp. 654–660. doi: 10.1038/s41586-022-04492-9.

Milborrow, S. (2015) ‘Notes on the earth package [WWW Document]’, URL http://www.milbo.org/doc/earth-notes.pdf, pp. 1–69.

Mutz, J., Roscoe, C. J. and Lewis, C. M. (2021) ‘Exploring health in the UK Biobank: associations with sociodemographic characteristics, psychosocial factors, lifestyle and environmental exposures’, BMC Medicine. doi: 10.1186/s12916-021-02097-z.

Peterson, R. A. (2021) ‘Finding Optimal Normalizing Transformations via bestNormalize’, R Journal. doi: 10.32614/rj-2021-041.

Sarris, J. et al. (2020) ‘Multiple lifestyle factors and depressed mood: a cross-sectional and longitudinal analysis of the UK Biobank (N = 84,860)’, BMC Medicine. doi: 10.1186/s12916-020-01813-5.

Schulz, R. et al. (2000) ‘Association between depression and mortality in older adults: The Cardiovascular Health study’, Archives of Internal Medicine. doi: 10.1001/archinte.160.12.1761.

Schwarzbach, M. et al. (2014) ‘Social relations and depression in late life - A systematic review’, International Journal of Geriatric Psychiatry. doi: 10.1002/gps.3971.

Smith, D. J. et al. (2013) ‘Prevalence and characteristics of probable major depression and bipolar disorder within UK Biobank: Cross-sectional study of 172,751 participants’, PLoS ONE, 8(11), pp. 1–7. doi: 10.1371/journal.pone.0075362.

Smith, S. M. et al. (2019) ‘Estimation of brain age delta from brain imaging’, NeuroImage. doi: 10.1016/j.neuroimage.2019.06.017.

Taylor, H. O. et al. (2018) ‘Social Isolation, Depression, and Psychological Distress Among Older Adults’, Journal of Aging and Health. doi: 10.1177/0898264316673511.

Terraneo, M. (2021) ‘The Effect of Material and Social Deprivation on Well-Being of Elderly in Europe’, International Journal of Health Services. doi: 10.1177/0020731420981856.

Townsend, P., Phillimore, P. and Beattie, A. (1988) Health and deprivation: Inequality and the North. Edited by P. Townsend, P. Phillimore, and A. Beattie. London: Croom Helm.

Yapp, E. et al. (2021) ‘Sex differences in experiences of multiple traumas and mental health problems in the UK Biobank cohort’, Social Psychiatry and Psychiatric Epidemiology. doi: 10.1007/s00127-021-02092-y.

Zhou, S. et al. (2021) ‘Socioeconomic status and depressive symptoms in older people with the mediation role of social support: A population-based longitudinal study’, International Journal of Methods in Psychiatric Research. doi: 10.1002/mpr.1894.

